# Impact of COVID-19 pandemic on cancer care delivery : A Real World Experience

**DOI:** 10.1101/2020.09.01.20183145

**Authors:** A Pandey, R Mala, C Neelam, P Mridula, S Ravindra, M Kanchan, Y Umesh, S Shivkant

**Affiliations:** Department of Medical Oncology, State Cancer Institute, Indira Gandhi Institute of Medical Sciences,Patna, Bihar, India

**Keywords:** COVID-19, chemotherapy, cancer care, Day care, lockdown

## Abstract

**Background:** There is lack of information on impact of Corona Virus Disease (COVID-19) pandemic on routine cancer care delivery.

**Aims and Objectives:** To evaluate the change in Day Care Chemotherapy (DCC) and Out Patient Department (OPD) patient numbers before and after COVID-19 national lockdown.

**Material and Methods:** Demographic data, diagnosis, type and frequency of chemotherapy delivered in Day Care between 1st February 2020 to 31st July 2020 were retrieved. Out Patient Department daily patient numbers were collected. Descriptive statistics, Odds ratio, Chi-square and Student T test were used to measure change in pattern of DDC and OPD patient numbers before and after 24th March 2020 (day of Lockdown). Pearson correlation coefficient was used to measure the strength of correlation between rise in COVID-19 cases and patient numbers.

**Results:** 3192 DCC and 8209 OPD visits were recorded in 126 working days. Median age was 47 years(SD + 19.06). Breast (17%) and Gall bladder(15%) were the most common cancers receiving chemotherapy. There was a significant decrease in number of DCC delivered in post COVID lockdown [mean 21.97 (+ 9.7)] compared to pre COVID lockdown [mean 33.30 (+11.4)], t=4.11, p = 0.001.There was a significant decrease in number of OPD visits in post COVID lockdown [mean 47.13 (+ 18.8)] compared to pre COVID lockdown [mean 89.91 (+30.0)], t=7.09, p = 0.001. The odds of receiving weekly chemotherapy over non weekly regimes significantly decreased post COVID lockdown with Odds ratio of 0.52 (95% CI, 0.36-0.75) with Chi square of 12.57, p =0.001. Daily COVID cases in State and OPD patient number were found to be moderately positively correlated on Pearson correlation coefficient, *r* = 0.35,p =0.001.

**Conclusion:** There was a significant fall in patient visit and chemotherapy cycles immediately after lockdown. The numbers increased later despite rise in COVID-19 cases.

## Introduction

On 24th March 2020, Government of India declared nationwide lockdown amidst fear of spurt in Corona Virus Disease (COVID-19) contagion.^1^ This decision was made to ‘flatten the curve’, delay the rise of COVID 19 related illness, buy time to improve heath infrastructure and build COVID care facilities. Many apex multi-speciality hospitals, including few cancer care delivery centres were designated as exclusive ‘COVID 19 facilities’.^2^ This rationing of services divested precious resources from other illness including cancer care. The problem was further compounded by intermittent disruption of care in running facilities whenever patients or healthcare workers turned COVID positive.

Strict lockdown with stringent travel restrictions often made patients afflicted with cancer to shun or delay opting of cancer care services, especially those residing in smaller towns and villages. This was further aggravated with concern of getting COVID, loss of wages, poor means to travel and reduction in healthcare workforce. This pose serious threat to upstaging of cancer, disease progression, symptomatic worsening including pain for patients who chose to stay home rather than avail therapy.^3^

Our hospital was declared as non COVID heath care facility and cancer treatment was available all through COVID 19 lockdown. In absence of paediatric oncology and haematology, department of medical oncology is the single point of contact for patients receiving chemotherapy irrespective of age and type (solid/ haematological) of malignancy in our centre. Recently, several expert opinion and review articles were published to rationalise and optimise cancer care delivery including risk stratifying patients in terms of urgency of treatment, delaying elective procedures, modifying chemotherapy to avoid toxicities, decreasing frequency to reduce hospital visits with minimal or no loss of overall efficacy and survival.^4-6^ We had the unique opportunity to study the trend of chemotherapy availed and compare the pattern before and after COVID lockdown. We report the first real world experience of impact of COVID 19 pandemic on routine cancer care delivery across several malignancies, before, during and after lockdown from India.

## Material and Methods

### General Study details

This is a retrospective case audit of number of patients visiting Out Patient Department (OPD) and subsequently receiving injectable chemotherapy in Day Care Centre (DCC) from 1^st^ February 2020 till 31^st^ July 2020. This study was conducted in the department of Medical Oncology in a single tertiary regional cancer centre, Indira Gandhi Institute of Medical Sciences (IGIMS), located in Eastern Indian State, Bihar. OPD and DCC services were available daily from Monday to Friday, except for Saturday, Sunday and days of national holiday. Written informed consent were obtained from all participants. The study was conducted according to the criteria mentioned in the International Conference on Harmonization Good Clinical Practices, Declaration of Helsinki, and guidelines established by the Indian Council of Medical Research. No funding was sought or received for this study. The Institutional Ethical Committee approval was duly obtained.

### Selection Criteria

All patients who received intravenous, intramuscular or subcutaneous injectable chemotherapy, irrespective of age and type of malignancy were entered in our database. Apart from chemotherapy, Granulocyte colony stimulating factor (G-CSF), injectable hormonal preparations, bone modifying agents, targeted therapy and immunotherapy, with or without chemotherapy were also eligible. Supportive care medications such as Non steroidal anti inflammatory drugs, opioids, antibiotics, anti-fungals, anti-emetics, crystalloids, colloids, iron preparations, if administered alone without concomitant chemotherapy were excluded. Any invasive procedure done in Day Care premises such as Peripheral inserted central catheter (PICC) insertion, flushing and dressing, ascitic or pleural effusion tapping, wound debridement and dressing were excluded from the study.

### Data Collection

Data was collected from records available in Day care chemotherapy (DCC) Unit attached to department of Medical Oncology. All cases registered, fulfilling selection criteria and received injectable chemotherapy on any working days from 1st February till 31st July 2020 were recorded. Demographic details including age, sex, type of malignancy, chemotherapy frequency, name of chemotherapeutic drugs, doses were retrieved and entered in database. Number of patients visiting in Out-Patient Department (OPD) of medical oncology during aforementioned working days were also obtained. The daily number of new COVID 19 cases detected in normal population in the State (Bihar) and State Capital City (Patna), where our institute is located was obtained from official Bihar government website/ twitter handle till date of 31^st^ July 2020.^7^

### Patient care and Health Care Worker (HCW) protection during COVID 19 pandemic

Our hospital was a designated non-COVID hospital by State Government and services including oncology were open and available throughout the study period. As per Institutional nodal Officer for COVID 19 and Hospital Infection Control Committee(HICC), Day Care and Out-Patient services of Medical Oncology Department was designated as ‘ moderate risk’ following Indian Council of Medical Research(ICMR) guidelines.^8^ While providing OPD consult and Day Care chemotherapy services, strict compliance to recommended level of Protective Personnel Equipments(PPE)- moderate risk as prescribed by ICMR advisory were followed.^9^ All doctors in OPD donned N95 mask, gloves, face shield and head cover; while nursing staff delivering chemotherapy donned all above in addition to shoe cover, goggles and overalls/gown.

All heath care workers attended regular training workshop for COVID 19 prevention as per recommendation by Institutional nodal officer and HICC. They were made aware of common signs and symptoms of COVID 19 and were instructed for prompt reporting in advent of above. If detected positive for COVID 19, the option for home / institutional quarantine or treatment if symptomatic was followed as per ICMR guidelines.^10^ At site of entry, HCW and patient/ relative were subjected to daily thermal screening. All except one able relative was permitted with patient while awaiting OPD and Day care services. All patients were asked to adorn N95 masks, while relatives were instructed to wear at least triple layered mask. The principles of social distancing and frequent hand washing or alcohol based hand sanitizer use was strongly encouraged

### Study scope and endpoints

I. The trend in number of patients who opted for Out-Patient Department and Day care chemotherapy services of Medical Oncology between 1^st^ February 2020 and 31^st^ July 2020. was evaluated.
II. The change in above case numbers were also measured and compared during following duration of pre-lockdown, serial nation-wide lockdowns, nation-wide unlocking and subsequent state-wide lockdown

A. Pre Lockdown/ routine/Baseline (1^st^ February 2020 till 24^th^ March 2020)
B. National Lockdown Phase 1 (25th March 2020 to 14th April 2020)
C. National Lockdown Phase 2 (15th April 2020 to 03th May 2020)
D. National Lockdown Phase 3 (04th May 2020 to 17th May 2020)
E. National Lockdown Phase 4 (18th May 2020 to 31st May 2020)
F. National Unlock Phase 1 (01st June 2020 to 30th June 2020)
G. National Unlock Phase 2 (01st July 2020 to 14th July 2020)
H. State Lockdown Phase 1 (15th July 31st July 2020)
III. Keeping the numbers of working days same (n=36), immediate change in OPD/ DCC patient numbers between Pre COVID 19 lockdown/ baseline (1st February 2020- 24th March 2020) and post COVID 19 lockdown (25th March 2020 - 15th May 2020) were compared. Changes in frequency or pattern of chemotherapy use, comparison of age group and chemotherapy regimens with respect to above two time points were done.
IV. As the pandemic escalated with time, there was rise in state wise and district wise COVID 19 cases in general population. At the same time, the national and state governments gradually deescalated lockdown and relaxed travel restrictions. We measured the change in trend of patients coming for cancer related services with actual rise of COVID 19 new cases in general population in later months (May, June, July 2020) of pandemic.

### Statistics

Descriptive statistics, frequency distribution, tables, charts and graphs were used to analyse the demographic, epidemiological, clinical and treatment related variables. Crosstabs, Odds ratio, Pearsons Chi-square test were used to compare binomial categorical variables. To compare means between pre and post lockdown case numbers, paired student t test was used. As the data was parametric, pearsons correlation coefficient was used to find association, its strength and direction between OPD/DCC patient numbers and rise in COVID 19 cases. Similarly, simple linear regression method was used to calculate relationship, if any between above two variables. All above statistics were computed and derived from SPSS software version 17.0 (IBM Corp. Released 2017. IBM SPSS Statistics for Windows, version 17.0. Armonk, NY, USA).

## Results

### Demographic, Clinical and Patient Profile

Out of 3238 registered cases in Day Care of Medical Oncology Department between 1^st^ February 2020 to 31^st^ July 2020, 3192 (98.57%) were eligible for this retrospective audit.(See Figure 1). During the aforesaid duration of six months, 8209 Out-patient Department (OPD) visits were registered over the total of 126 working days. Median age of population receiving treatment in Day care was 47 years(SD + 19.6) with male to female ratio of 1.10. Breast cancer 543 (17%) and Gall Bladder cancer 488 (15.2%) were the most common malignancies seen. Three weekly chemotherapy was the most commonly prescribed schedule with 1990 cycles (62.4%). The most common age group receiving chemotherapy was adults (age 18-60 years) with 2101 cases (65.8%). Chemotherapy with or without targeted therapy was the most common therapy received in Day care with 3000 cycles (93.9%). See Table 1.

**Figure 1.**
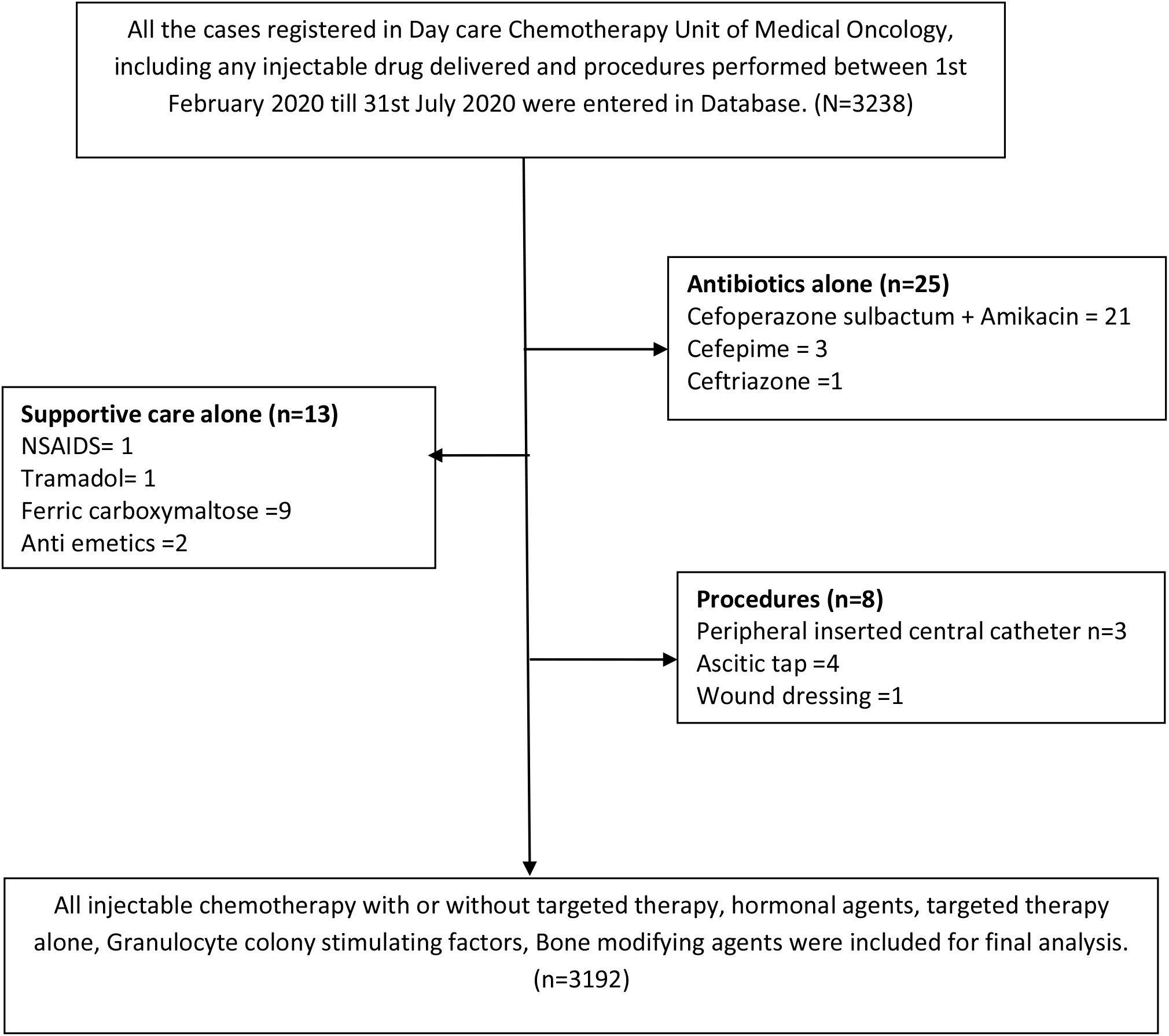
The flow diagram showing the number of cases who received various treatment in our Medical Oncology Day care and selection algorithm of final cases who were enrolled in this study

**Table 1.** Demographic, disease and treatment related variables of patients who received injectable drugs / cycles between 1^st^ February 2020 to 31^st^ July 2020.

| Sr. No | Variables | Value/ Numbers | Remark/Percentage |
| --- | --- | --- | --- |
| 1 | **Age** | Median 47 years ( ± 19.06) | Range 87 (1-88) |
| 2 | **Sex** |  |  |
|  | Male | 1677 | Ratio 1.10 |
|  | Female | 1515 |  |
| 3 | **Age Group** |  |  |
|  | Paediatric (Age 1-18 years) | 454 | 14.2% |
|  | Adult (18-60 years) | 2101 | 65.8% |
|  | Elderly (above 60 years) | 637 | 20.0% |
| 4 | **Chemotherapy Frequency** |  |  |
|  | Weekly | 327 | 10.2% |
|  | Two weekly | 483 | 15.1% |
|  | Three weekly | 1990 | 62.4% |
|  | Four weekly | 392 | 12.3% |
| 5 | **Type of Drugs administered** |  |  |
|  | Chemotherapy alone | 2702 | 84.6% |
|  | Chemotherapy+ targeted | 298 | 9.3% |
|  | Targeted alone | 101 | 3.2% |
|  | Growth Factors | 39 | 1.2% |
|  | Hormonal Agents | 20 | 0.6% |
|  | Bone modifying agents | 18 | 0.6% |
|  | Immunotherapy | 11 | 0.3% |
|  | Immunotherapy+Chemotherapy | 3 | 0.1% |
| 4 | **Type of cancers** |  |  |
|  | Breast | 543 | 17.0% |
|  | Gall Bladder | 488 | 15.2% |
|  | Lung | 386 | 12.1% |
|  | Ovary | 251 | 7.9% |
|  | Colo-rectal | 245 | 7.7% |
|  | Acute Lymphoblastic Leukaemia | 222 | 7.0% |
|  | Non Hodgkins Lymphoma | 118 | 3.7% |
|  | Upper Gastrointestinal | 117 | 3.7% |
|  | Multiple Myeloma | 115 | 3.6% |
|  | Uro-genital | 113 | 3.5% |
|  | Wllms tumour | 89 | 2.8% |
|  | Pancreas | 75 | 2.3% |
|  | Hodgkins Lymphoma | 74 | 2.3% |
|  | Head neck | 53 | 1.7% |
|  | Ewings Sarcoma | 50 | 1.6% |
|  | Others | 253 | 7.9% |

### Trends with respect to time and lockdown

Month wise consecutive changes in patient numbers receiving therapy in Day Care is shown in Table 2. Change in demographic profile, including age group and frequency of chemotherapy regimens delivered also varied between February 2020 and July 2020 (See Table 2). Median number of Out -Patient Department (OPD) and Day Care chemotherapy (DCC) cases from pre-lockdown baseline dropped by 66.66% and 53.85 % respectively immediately following imposition of nation-wide lockdown. (See table 3) After gradual recovery in number of cases in later months (May-June 2020) with reduction in travel restrictions and unlocking, the case numbers plummeted again after 15th July 2020 onwards due to imposition of state-wide lockdown.(See Figure 2A and 2B)

**Table 2.** Change in trend of age groups, frequency of drugs administered and type of drugs between 1^st^ February 2020 to 31^st^ July 2020.

| Sr. No | Groups | February 2020 (%) | March 2020 (%) | April 2020 (%) | May 2020 (%) | June 2020 (%) | July 2020 (%) |
| --- | --- | --- | --- | --- | --- | --- | --- |
| 1 | **Age groups** |  |  |  |  |  |  |
|  | Paediatric | 58 (8.7%) | 46 (8.3) | 30 (6.8) | 77 (17.8) | 117(21.0) | 134(24.8) |
|  | Adult | 455 (68.6%) | 393 (70.4) | 309 (70.5) | 269 (62.5) | 338 (60.3) | 333(61.7) |
|  | Elderly | 151 (22.7%) | 119 (21.3) | 100 (22.7) | 85 (19.7) | 105 (18.7) | 73(13.5) |
| 2 | **Frequency** |  |  |  |  |  |  |
|  | Weekly | 84(12,6) | 46(8.3) | 18(4.1) | 51(11.8) | 66(11.8) | 62(11.5) |
|  | Two weekly | 111(16.7) | 103(18.4) | 74(16.8) | 52(12.0) | 67(11.9) | 76(14.0) |
|  | Three weekly | 391(59.0) | 333(59.7) | 296(67.5) | 280(65.1) | 366(65.4) | 324(60.0) |
|  | Four weekly | 78(11.7) | 76(13.6) | 51(11.6) | 48(11.1) | 61(10.9) | 78(14.5) |
| 3 | **Type of drugs** |  |  |  |  |  |  |
|  | Chemotherapy alone | 472(84.7) | 375(85.6) | 548(82.6) | 366(85.0) | 477(85.1) | 464(86.0) |
|  | Chemotherapy and targeted therapy | 46(8.3) | 38(8.6) | 74(11.2) | 37(8.6) | 49(8.7) | 54(10.0) |
|  | Targeted therapy alone | 20(3.6) | 12(2.7) | 27(4.1) | 11(2.5) | 19(3.4) | 12(2.2) |
|  | Growth factors | 6(1.0) | 9(2.0) | 5(0.7) | 8(1.9) | 7(1.3) | 4(0.7) |
|  | Bone modifying agents | 5(0.9) | 3(0.7) | 3(0.4) | 4(0.9) | 2(0.4) | 1(0.2) |
|  | Hormonal | 2(0.3) | 1(0.2) | 4(0.6) | 4(0.9) | 4(0.7) | 5(0.9) |
|  | Immunotherapy | 6(1.1) | 1(0.2) | 3(0.4) | 1(0.2) | 0(0.0) | 0(0.0) |
|  | Immunotherapy and chemotherapy | 1(0.1) | 0(0.0) | 0(0.0) | 0(0.0) | 2(0.4) | 0(0.0) |

**Table 3.** Median number of Out Patient Department and Day care Chemotherapy in several sequential phases of national lockdown, unlocking and state lockdown with change from baseline (pre-lockdown) values between 1^st^ February 2020 to 31^st^ July 2020.

| Sr. No | Time Points | Working days (n) | Out Patient Department |  | Day care Chemotherapy |  |
| --- | --- | --- | --- | --- | --- | --- |
|  |  |  | Median cases (+/- SD) | Change from Baseline | Median cases (+/- SD) | Change from Baseline |
| 1 | Pre LOCKDOWN/<br>routine/Baseline<br>OPD | 35 | 99 ( + 2.7)<br>115 (31-146) | NA | 39 ( + 1.14)<br>42 (11-53) | NA |
| 2 | National<br>LOCKDOWN Ph 1<br>(25-03-2020 to 14-4-2020) | 16 | 33 ( + 2.18)<br>81 (6-87) | - 66.66% | 18 ( + 6.79)<br>22 (6-28) | - 53.85% |
| 3 | National<br>LOCKDOWN Ph 2<br>(15-04-2020 to 03-05-2020) | 12 | 56 ( + 9.9)<br>36 (45-81) | - 43.44% | 23 ( + 9.04)<br>32 (12-44) | - 41.1% |
| 4 | National<br>LOCKDOWN Ph 3<br>(04-05-2020 to 17-05-2020) | 9 | 54 ( + 1.07)<br>31 (35-66) | - 45.46% | 26 ( + 1.13)<br>40 (12-52) | - 33.34% |
| 4 | National<br>LOCKDOWN Ph 4<br>(18-05-2020 to 31-05-2020) | 9 | 53 ( + 1.31)<br>38 (32-70) | - 46.47% | 24 ( + 8.3)<br>25 (13-38) | - 38.47% |
| 5 | National UNLOCK<br>Ph 1 (01-06-2020 to 30-06-2020) | 22 | 65 ( + 9.97)<br>33 (51-84) | - 34.35% | 24.5 ( + 7.8)<br>32 (11-43) | - 37.18% |
| 6 | National UNLOCK<br>Ph 2 (01-07-2020 to 14-07-2020) | 10 | 76 ( + 1.57)<br>44 (44-88) | - 23.24% | 25 ( + 8.26)<br>24 (17-41) | - 35.90% |
| 7 | State Lockdown Ph 1<br>(15-07-31-07-2020) | 13 | 53 ( + 1.26)<br>42 (24-66) | - 46.47% | 23 ( + 5.81)<br>21 (11-32) | - 41.03% |

**Figure 2A and 2B.**
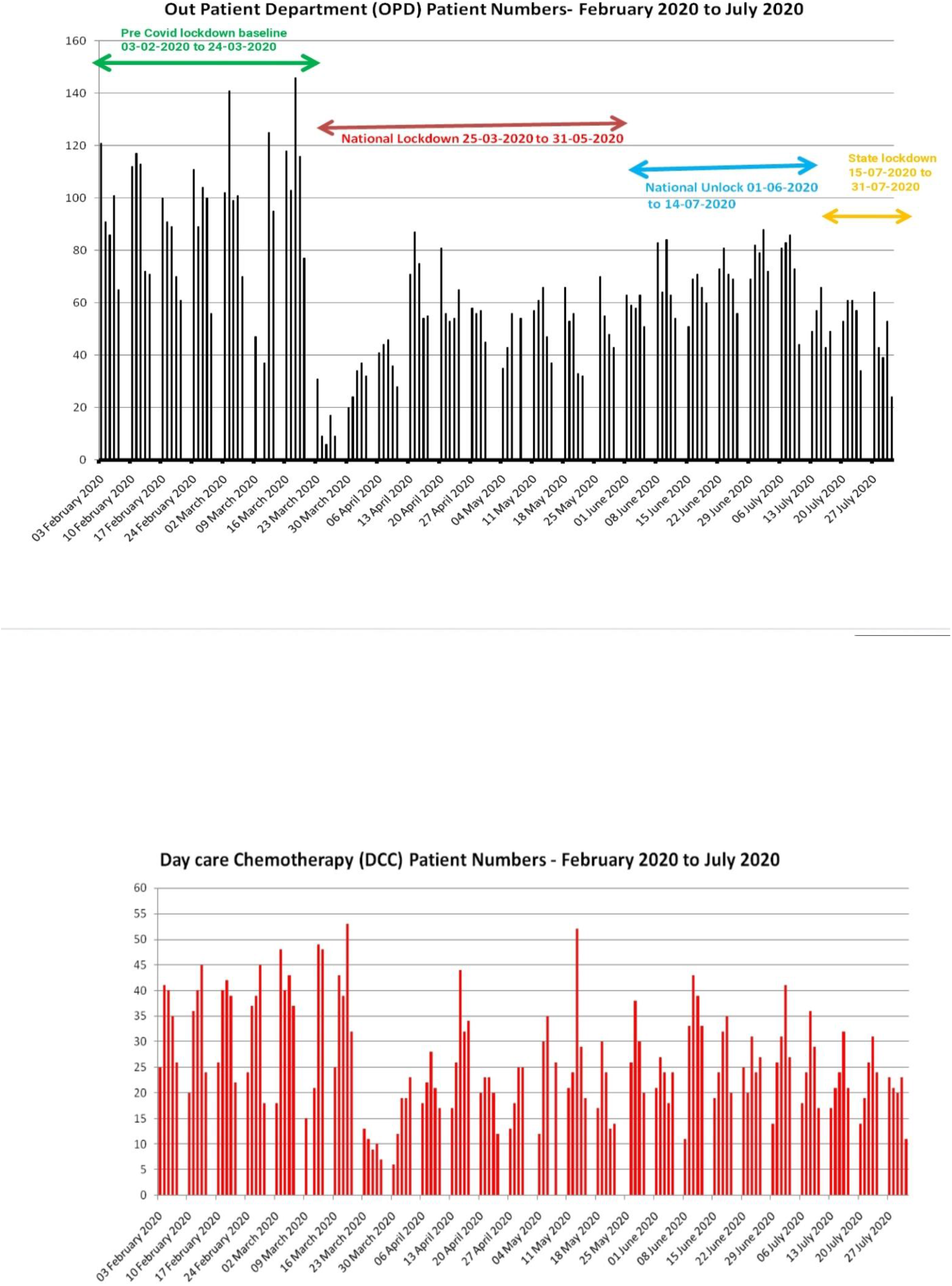
Out-patient Department (OPD, Figure 2A) and Day care Chemotherapy (DCC, Figure 2B) trend between 1^st^ February 2020 to 31^st^ July 2020 in our Medical Oncology Department.

### Comparison between Pre COVID baseline and post COVID 19 lockdown

Keeping the number of working days same (n=36) in both groups for homogeneity, namely Pre COVID lockdown(1st February 2020- 24th March 2020) and Post COVID lockdown (25th March 2020 - 15th May 2020), DCC and OPD were compared using paired student t test. There was a significant decrease in number of DCC delivered in post COVID lockdown [mean 21.97 (+ 9.7)] compared to pre COVID lockdown [mean 33.30 (+11.4)], t=4.11, p = 0.001.Similarly, there was a significant decrease in number of OPD visits in post COVID lockdown [mean 47.13 (+ 18.8)] compared to pre COVID lockdown [mean 89.91 (+30.0)], t=7.09, p = 0.001. There was more proportionate drop in weekly chemotherapy cycles from 128 to 41 (68%) than that of three weekly, 703 to 469 (33%) when compared between pre COVID and post COVID lockdown, suggesting preference to switch to regimens with higher intervals.

The odds of receiving weekly chemotherapy over non weekly regimes significantly decreased post COVID lockdown with Odds ratio of 0.52 (95% CI, 0.36-0.75) with Chi square of 12.57, p =0.001. Among patients who received paclitaxel alone or paclitaxel based chemotherapy, the odds of receiving weekly paclitaxel decreased compared to three weekly paclitaxel post lockdown by Odds ratio of 0.10 (95% CI0.03-0.29) with Pearsons Chi square of 22.68, p =0.001. Among patients of colorectal cancer receiving biweekly (FOLFOX/FOLFIRI) or three weekly (CAPOX/CAPIRI) based regimes, biweekly regimes showed a non significant decreased trend post lockdown, with Odds ratio of 0.58 (85% CI- 0.23-1.45).Paediatric and elderly population had odds ratio of 1.92 (95% CI, 0.86-1.60) and 0. 93 (95% CI, 0.74-1.17) respectively for receiving chemotherapy in post COVID lockdown versus pre COVID lockdown which were not significant.

### Change in trend of patients opting for cancer treatment with rise of COVID 19 cases in general population

Though the nation-wide lockdown was imposed on 24th March 2020 to curb COVID 19 pandemic, the actual rise of new COVID 19 cases in our State began in late April and early May. As the cases of COVID 19 cases continue to surge ahead, gradual de-escalation, unlocking and reduction in travel restrictions was allowed in graded manner (See Figure 3A, 3B). Daily COVID cases in State (Bihar) and Outpatient Department patient number were found to be moderately positively correlated on Pearson correlation coefficient, *r* = 0.35, p =0.001; but not for city (Patna) or DCC. With positive Pearson correlation, Linear regression was used to access whether rise in new COVID cases predict change in Outpatient patient(OPD) numbers. The linear regression model was significant with F =12.39, p=0.001. Number of OPD cases could be predicted from rise in new COVID cases in State by the following formula: OPD cases= 1.724 x New COVID cases - 6.704, R square =0.123.

**Figure 3A and 3B.**
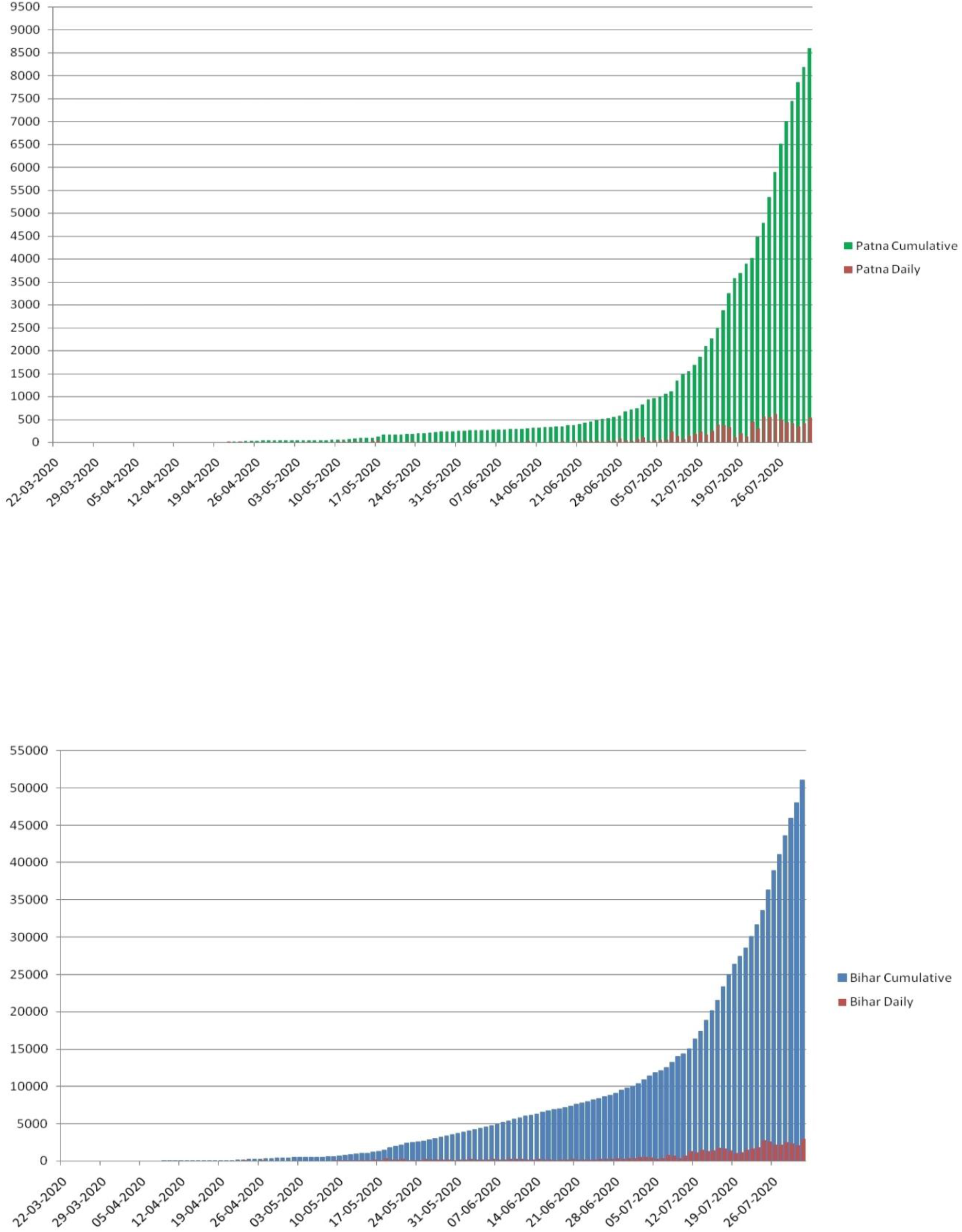
Daily new and cumulative COVID 19 cases detected in City (Patna, Figure 3A) and State (Bihar, Figure 3B) where our institute in located with time.

## Discussion

COVID 19 pandemic and efforts to curb its spread affected routine cancer care delivery. Nation-wide lockdown, travel restrictions and fear of getting infected with COVID caused treatment delay and reduction in number of patients availing cancer directed services. In our study, there was a significant decline in number of Out Patient Department (OPD) visits immediately post lockdown [mean 47.13 (± 18.8)] compared to pre COVID lockdown [mean 89.91 (+30.0)], p = 0.001. There was also fall in number of Day Care Chemotherapy (DCC) delivered in post COVID lockdown [mean 21.97 (± 9.7)] compared to pre COVID lockdown [mean 33.30 (±11.4)], p = 0.001. We also noticed change in pattern of prescription with significant decrease in weekly chemotherapy cycles as pandemic progressed, with intention to decrease patient visit frequency, Odds ratio of 0.52 (95% CI, 0.36-0.75) with Chi square of 12.57, p =0.001. However, as the lockdown restrictions were relaxed, the patient numbers in OPD increased despite rise in COVID 19 cases in community in later months, suggesting subduing of COVID concerns with cancer therapy delay implications and disease progression.

A survey done on clinicians practising gynaecologic oncology found significant fall in patient volumes, especially in government hospitals after COVID lockdown.^11^ In an early analysis reported by Patil et al. showed drop in footfalls in a retrospective audit of adult medical head neck cancer unit of apex tertiary cancer hospital.^12^ Indian Association of Surgical Oncology mandated routine COVID-19 testing before any surgery.^13^ Patients for curative cancer surgeries were postponed if found to be COVID positive and referred for neoadjuvant chemotherapy, radiotherapy, hormonal therapy or sometimes best supportive care till viral clearance or recovery of health status.^14-16^ Our study portrays the real world experience over the time period of six months of running Out Patient Department and Day Care chemotherapy services during COVID pandemic. We demonstrate significant decline in patient numbers immediately post lockdown. We also found significant reduction in prescriptions of weekly chemotherapy post lockdown. One of the prospective study conducted in Oncology patients demonstrated the willingness to continue chemotherapy during the pandemic, as they perceived disease progression as bigger threat than COVID.^17^ Our study substantiate their claim, as immediately after lockdown restrictions were lifted, our Out-Patient Department patient numbers started rising despite rise in COVID 19 cases in general population, which had moderately positive correlation to it on Pearsons correlation coefficient.

Delay or postponement of radiotherapy or systemic therapy affects cancer outcomes adversely in head neck, colon and breast cancer, especially in curative setting.^18-20^ Moreover, patients diagnosed with acute leukaemia, high grade lymphoma, metastatic germ cell tumour require urgent intervention with systemic therapy to prevent mortality. Several expert opinions and recommendations were issued regarding approach to cancer patients during COVID 19 pandemic. ^3-6,21^ Guarded approach to systemic therapy with prompt intervention for urgent medical crisis and tumours with curative potential, while delay in palliative chemotherapy for cases with limited or questionable survival benefit was advocated in most. Our cancer centre is within a tertiary government multi-speciality hospital. During lockdown, all patients were initially screened at ‘flu centre’ and then referred to OPD consult or chemotherapy, as practised by another apex cancer centre.^22^ However, we did not routinely tested our patients for COVID 19 before OPD consult or chemotherapy unless symptomatic of COVID as per ICMR recommendation.^23,24^ None of our healthcare workers during the study period were tested positive for COVID-19, reported sick or absent. Neither Out Patient Department nor Day care services were interrupted on any working days during aforesaid period.

Our study did not have details of patients except their numbers who visited Outpatient Department(OPD) unlike those receiving chemotherapy in Day care. Hence, variation in type of patients in OPD, type of cancer, oral chemotherapy done on OPD basis or any change from intravenous to oral therapy during lockdown are unavailable. We also do not have information on whether any maintenance chemotherapy such as pemetrexed in lung cancer or 5- Fluorouracil in colon cancer were halted, increased in frequency or changed to oral therapy during lockdown as this decision was often made in OPD. We also do not have information on any of our cancer patient who turned COVID positive in any other centre or COVID designated hospital, abandoned therapy or died due to it. New patients otherwise eligible for curative surgeries or radiotherapy were directed to us for neo adjuvant chemotherapy or hormonal therapy as elective surgery was postponed during lockdown. We do not have separate information on those cases. Being retrospective, Day Care Chemotherapy(DCC) register did not mentioned stage and intent of therapy, hence this information in missing in our analysis. As none of the asymptomatic healthcare workers were tested for COVID 19, any asymptomatic COVID disease or carrier state in them is unknown. As many of our patients braved travel restrictions during lockdown and came from far off places, we did not deny them chemotherapy even if less urgent or for minimal symptomatic palliative benefit.

## Conclusion

Nationwide lockdown to curb COVID 19 pandemic had negative impact on cancer treatment with significant reduction in number of patients opting for cancer directed therapy. The pattern of chemotherapy prescriptions changed to larger interval and protracted course to reduce frequent patient visits. Withdrawal of travel restrictions later led to rise in patient numbers for consultation despite rise of COVID 19 new cases detected in community.

## Data Availability

Raw data were generated at our Institute (Indira Gandhi Institute of Medical Sciences). Derived data supporting the findings of this study are available from the corresponding author [initials] on request.

## Acknowledgements

None

